# Reproduction Number EXCEL Model of COVID-19 for predictive calculation of nearby trend up to the end of infection

**DOI:** 10.1101/2023.08.11.23293983

**Authors:** Shinzaburo Matsuda, Hiroshi Toki

## Abstract

Reproduction Number (RN-)EXCEL Model has been developed on an EXCEL sheet to provide important characteristics of COVID-19 infectious disease for practical use. The model is developed based only on observed data to predict future infection toward herd immunity threshold and until the end stage of the infection. Basic equations are simple and constructed in analogy with neutron multiplication reactions in nuclear reactor. To know the next day infection, we calculate an exponential increase in one day step with a rate obtained from nearby PCR positive infectious numbers, which are daily input in the EXCEL sheet. In a closed community, main players are non-immune holders and immune holders, where total number of immune holders derived from infection and vaccination plays an essential role. In traditional SIR model, infection behavior is characterized by the reproduction rate in differential equation where social actions such as governmental regulations or vaccinations are included as constant breaking term for infection spread. However, in actual situation these terms are time dependent and is difficult to solve by a set of differential equations. In contrast, RN-EXCEL model deals with infection by defining successive reproduction number for each time interval as inchworm that represents a clear physical picture of the virus infectivity. Using this model, a lot of predictions were made for semi-closed communities domestically and world-wide, timely for practical use.

## 1. Introduction

Coronavirus disease 19 (COVID-19) and a pandemic was declared by the World Health Organization (WHO) in 2019. COVID-19 has been impacting a large number of people worldwide, and as of 2020/4/7, around 1,400,000 cases worldwide have been reported [1]. About 8% of world population have been infected by the time of the beginning of 2023 [2]. There are many papers published on COVIT-19, which have been influencing daily life worldwide. Instead of referring many papers here, we list some publications, which influenced the present study.

To describe the development of the infectious disease, it is essential to understand the mechanism of successive infection and herd immunity. In an infectious disease, an infected person infects *m* persons during his/her infected period. This is the first generation, and the same processes are continued for the successive generations. Here, *m* represents infectivity, and the number of infected persons increases exponentially. As time goes, there are substantial amount of immune holders, which decreases the number of non-immune holders to be infected. Eventually a community arrives at a point called herd immunity, where infectious disease stops increasing. We tried to emphasize this physical picture and developed a simple mathematical form by index function. This form together with daily input of the number of infected persons [2, 3] are key variables in an EXCEL sheet and provide an overview of the COVID-19 infection. It is mathematically equivalent to solve difference equations suitable for the data given by daily variables. In the early phase of infection, both SIR model and our RN-EXCEL model provided essentially the same results for prediction, including herd immunity [4, 5].

With a progress of infection, breaking factor such as governmental regulation and behavior change of public are necessary to be considered [2, 6]. Since these factors are time dependent, however, they cannot be included in one set of differential equations in the SIR model. Therefore, a series of equations for appropriate time intervals are necessary and their solutions must be interconnected. In contrast, RN-EXCEL model deals with difference equation from the beginning and can avoid these difficulties by introducing reproduction number *m*(*t*) in which all of these factors are reflected. Namely, *m*(*t*) for each time interval is an important measure of infectivity between competing SARS-CoV-2 virus and resident people. When the corona vaccination started near the end of 2020, we took the vaccination data including their effectiveness into calculation as breaking term for infection spread [7, 8, 9]. This made a straightforward change on the number of immunity holders.

Another important fact of COVID-19 was the existence of asymptomatic and mild infected people that escaped PCR tests. A ratio of these people to the PCR test positive infected people can be estimated by the combination of public antibody test and cumulative infection numbers [7, 8, 9, 10]. Presence of a large unidentified infectious persons made difficult for the governmental organization to follow infection routes and to make proper isolation measure. Asymptomatic infected persons dropped vaccination efficiency targeting the non-immunity holders. Therefore, repetitious antibody tests for public are indispensable at proper time intervals and proper size for estimation of immune holders and prediction.

At the latest phase of infection approaching herd immunity conditions, some countries face to reinfection [11, 12, 13]. In the RN-EXCEL model, without taking decay of immunity effectiveness of infected persons in the background of reinfection into calculation, the calculation could not follow observed infected numbers [14]. Taking all these changes, we could study COVID-19 from the beginning continuously up to now. It deeply depends on RN-EXCEL model that accepts daily changing corona data quite efficiently. We used this RN-EXCEL model for calculation in many countries and prefectures and found its effectiveness for actual use. Therefore, not only for prediction of the ongoing COVID-19 infection, we believe this method is useful and is used world-wide for future infections. It is our hope that scientific staffs to be engaged in future infection works could be prepared to use this model in advance of the next infection.

## 2. Reproduction Number EXCEL Model

### 2.1. Fundamentals of RN-EXCEL model

The RN-EXCEL model deals with a closed community where fixed total number of population *A* is composed of either non-immune holders or immune holders as illustrated in Fig. 1. A part of the non-immune holders *B*(*t*) are infected and then infects other persons during their active *τ* days but eventually resurged and get immunity. Namely, they now are converted to an immune holder *R*(*t*) and are not subject to infection again. Compared to the size of these two state variables, the number of persons currently under infection in hospital or in house is much smaller and is neglected in modeling the RN-EXCEL model. Likewise, the number of deceased persons are small and also neglected because of its smallness as compared to *B*, and *R*. Hence, we write *B*(*t*) + *R*(*t*) = *A*. In addition to this basic route to get immunity, two other routes should be considered. One is vaccination, and the other is asymptomatic infection.

**Figure 1:**
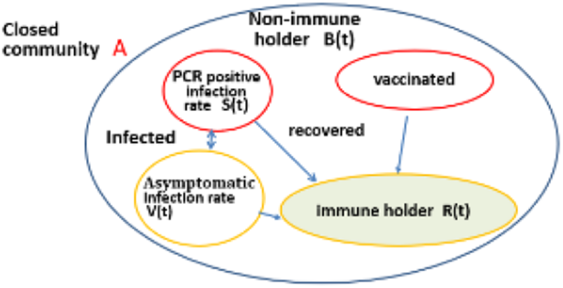
The structure of infection and immunity for a closed community with the number of inhabitant *A*.

For the construction of the present model, we assume first that one infected person infects *m* persons during active period of *τ* days, where *m* is a multiplication factor and *τ* is a cycle time of one generation of infection. So, at the end of 2nd generation (2*τ* days later), *m*^2^ persons are infected. This leads to 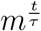 persons are infected after *t* days. Thus, the number of outbreak of infection at time *t, S*(*t*) is expressed as

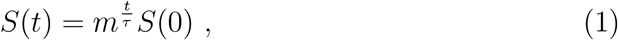

where *S*(0) is an initial value at *t* = 0. Choosing one day for the time step, daily infected persons at the *j*-th day is then expresses as,

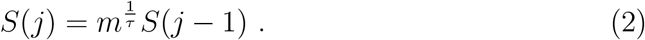

Thus, with daily inputs of the number of infected persons, we obtain a set of (*m, τ*). We now have a freedom of determining *m* and *τ*, but choose 10 days for *τ* by clinical experience of an active infection period until recovery. Then we determine *m* that has a clear physical picture of infectivity of the virus concerned.

People once infected and recovered could hold acquired immunity, a basis of the herd immunity concept. When infection expands and more persons hold immune, an infected person can only find smaller number of susceptible non-immune holders so that infection efficiency is decreased. This is illustrated in Fig. 2 for a case of *m* = 3 at the time when 40% of the population has already immunity. Even the infected person (red) may closely contact three persons, only 60% of the population are susceptible. Therefore, one infectious person makes reduced infectivity but still an effective reproduction number is 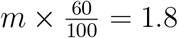 which is larger than 1.0. Namely, one infected person still generates 1.8 infected persons and that means infection spreads. In addition to the infection immunity, vaccination also generates immune holders. Thus, the probability of effective infection is decreased by the presence of immune holders. Namely, infectivity is expressed by the effective reproduction number as,

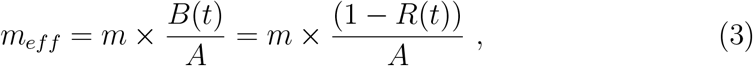

and if non-immune holders are reduced to 1/3 (33.3%) of the population, *m* is effectively reduced to 1.0, i.e. *m*_*eff*_ = 1.0, the herd immunity threshold is achieved. The reduction factor *B*(*t*)*/A* is regarded as shielding effect by the immune holders, and we replace *m* in equation (2) by *m*_*eff*_ to yield,

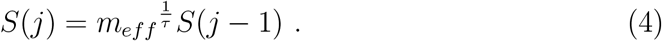

Infection decline or spread depends on *m*_*eff*_ *≤*1 or *m*_*eff*_ *≥* 1, which is easily seen from Eq. (4). Up to this point, our model is almost the same as the SIR model. Differences are the virus infectivity *m* and the total immune holder *R*(*t*) that are changing quite frequently. These variables are always monitored for predictive calculations in our model, while in the SIR model the multiplication rate should be used and to be changed frequently as explained in the Appendix.

**Figure 2:**
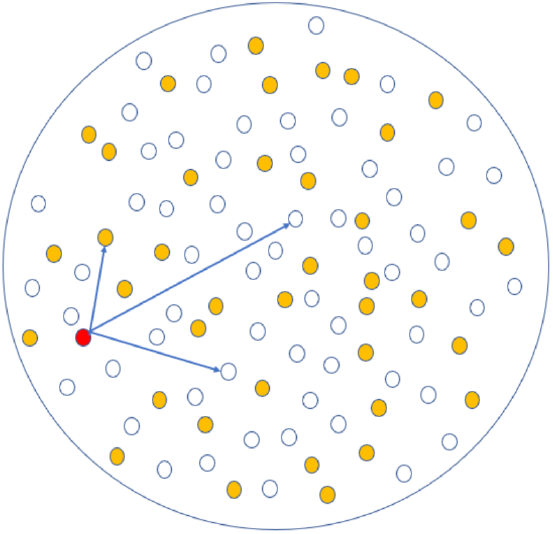
Illustration of infection by the active infectious person (red) for the case of *m*=3 at the time when 40% of population already have immunity. The active person (red) may closely contact three persons shown by arrows, but 40% of the population (orange color) already have immunity and are not infected, and 60% of the population (white) are non-immune holders and subject to infection.

One important point is that, suppose virus is strong like *m* = 10, then herd immunity threshold *m*_*eff*_ = 1.0 is achieved from Eq. (4), if

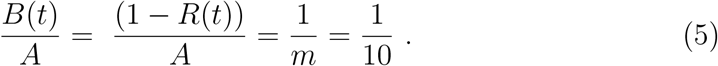

Namely, non-immunity holder is less than 10% of the population, and the immune holders is as high as 90%.

It is interesting to mention that the infection process is analogous to a simplified form of the thermal neutron multiplication in the light water reactors. The control rod or Boron as neutron absorber controls the number of thermal neutrons for their multiplication [15] just like the immune holder in COVID-19. The reactor critical condition corresponds to the herd immunity threshold.

### 2.2. Definition of Variables, Constants, and Parameters

In an EXCEL sheet, typical inputs in each column are: *t*: date of event, taking one day step, *t* is expressed as an index *j*.

*S*(*t*): number of persons observed PCR test positive (persons per day), and averaged over seven days.

*S*_*cal*_(*t*), and *S*_*int*_(*t*): calculated infection number of persons, and its cumulative number.

*V*_*cal*_(*t*), and *V*_*int*_(*t*): calculated asymptomatic infection rate, and its cumulative number.

*R*(*t*): number of immune holders.

*B*(*t*): number of non-immune holders.

*Q*(*t*), and *P* (*t*): cumulative vaccination numbers, and its effective immune holders.

*μ*(*t*): the effectiveness of the vaccination

*α*(*t*): a ratio of asymptomatic infected persons to PCR positive infection persons.

Constants: *A, τ*, time step Δ*t*, and parameters of decay of the vaccine effectiveness.

We write here how to get the ratio *α* of asymptomatic infected persons to PCR positive infection persons. Since the time step of one day is adopted, *t* is replaced by *j*, and the cumulative numbers at the *j*-th day from the beginning are written as

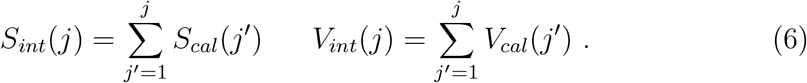

At time *t*, which is indexed as *j*, we know the ratio of *S*_*int*_(*j*)*/A* for some community with population *A* by accumulating the daily information. If antibody tests are conducted in the same community, but in a reduced scale (*A*_*s*_ is a sample number) in the community, a ratio of persons once infected (*S*_*int*_(*j*) + *V*_*int*_(*j*))*/A*_*s*_ is known. We assume this ratio scales to the total community population, as long as sufficient numbers of test donors are selected without bias. Then, (*S*_*int*_(*j*) + *V*_*int*_(*j*))*/A* is known, and as a result, a ratio of *V*_*int*_(*j*)*/S*_*int*_(*j*) = *α*(*j*) is derived.

Meaning of *α* is significant because asymptomatic infection generates 1+*α* times larger immune holders as compared with the familiar PCR positive infection immune holders. In the first year of COVID-19, antibody tests in each country provided a high number, more than 10 in NY states [10], cities in USA, UK, India, South Africa, and more than 5 in Japan and many other European countries. Since *α* is time dependent, we take every data at a time of antibody tests conducted in each country, calculate *α* and make interpolation between the data to get *α*(*t*). Values of *α*(*t*) generally tends to decrease with time and in one or two years later it decreased to about 1 *∼* 2 or less depending on the country with a very low changing speed. After the latest antibody test, we use extrapolation of the *α*(*t*) curve. For the prediction purpose, *α* is very important to estimate the number of PCR positive infected persons from two aspects. First, in nearby prediction, for a fixed *m* value, the peak of infection will arrive earlier with the presence of asymptomatic infected persons, because the non-immunity holders decrease much faster than the case with only PCR infections. Second, distance between the present immunity rate and the herd immunity threshold can be bridged with a total number of PCR positive and asymptomatic infections.

With the active time of *τ* days, a cumulative number *R*(*j*) of immune holders can be expressed by using the number of daily infected persons *S*_*cal*_ including the contribution from vaccination *P* (*j*) as

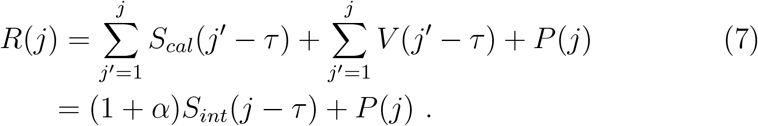

While systematic data for vaccine effectiveness is limited, UK COVID-19 vaccine surveillance report (week 16: 2022/4/16) provided updated data on the decay of vaccine effectiveness against infection by the initial two doses and recovery by the successive 3rd booster vaccination [9]. The data provided the vaccine effectiveness and its decay for various combinations of vaccines. In this report, vaccine effectiveness for omicron variant is decaying from about 75% just after the injection down to 25% level after 20-24 weeks, which is much faster than for the delta variant. When 3rd booster vaccine is injected, the effectiveness recovers but not fully, and it also decays from the 3rd booster time. We emphasize that the decay of the effectiveness means some persons, who once get vaccine immunity, lose immunity against infection and return to the non-immunity group.

To take these data into model calculation is somewhat complicated. Vaccination data are generally obtained by the cumulative number as a function of date. The difficulty of vaccine is the gradual decay of the effectiveness against infection, which depends on the numbers and combinations of vaccine injections and further COVID-19 strain types. All these informations are not generally provided at the daily basis. Hence, we have to take a heuristic implementation of the vaccine effect.

As for the number of injection per day, we take the increment Δ*Q*(*j*) = *Q*(*j*) *− Q*(*j −* 1). Using this number, we can calculate the immune holders by vaccination by multiplying the decay function *μ*(*j*) to the increment Δ*Q*(*j*) as

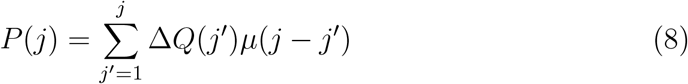

This implementation is numerically cumbersome, and further the informations of injection and the decay rates for various virus strain types for sufficient time period are not available. The decay curve for *μ* after injection seems to drop rapidly for instance for omicron until 20-24 weeks after the third injection and stays almost constant [9]. Here, we use *μ*(*j − j*^*′*^ *>* 154) = *μ*_0_ = *μ*(*j − j*^*′*^ = 154), the decayed effectiveness at 154th days after injection, the center value of 20-24 week. Hence, we divide the above equation into two terms:

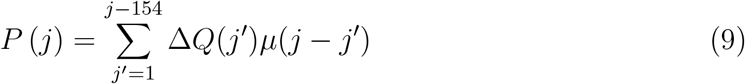

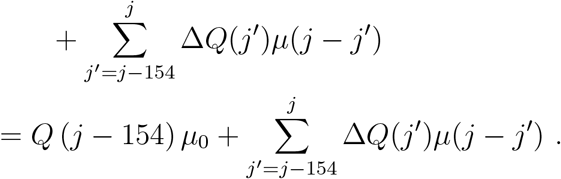

The contribution of the second term is calculated for a few cases explicitly and estimated to be about 10% of the first term. Eventually *P* (*j*) is approximated as,

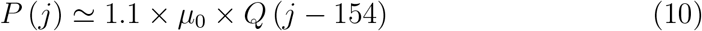

This simplification gives a practical lower estimate of the influence of the past vaccination.

### 2.3. How to determine m from the observed infection rate

Now we explain how we study a feature of the COVID-19 virus from the observed data [2]. Fig. 3 shows how to determine the reproduction number *m* from the best fit curve. Near the end of the infection wave, suppose that the best fit curve for the time period between A and B is obtained as shown by a red curve with a reproduction number of 0.9, the observed infection data beyond B is deviating from this curve. Then, setting B as starting point and by changing *m*, the best fit curve is obtained as dotted red curve with a new *m* value of 1.1 and after fitting, *S*(*t*) = *S*_*cal*_(*t*). We name this as inchworm method to determine *m* within a certain period successively.

**Figure 3:**
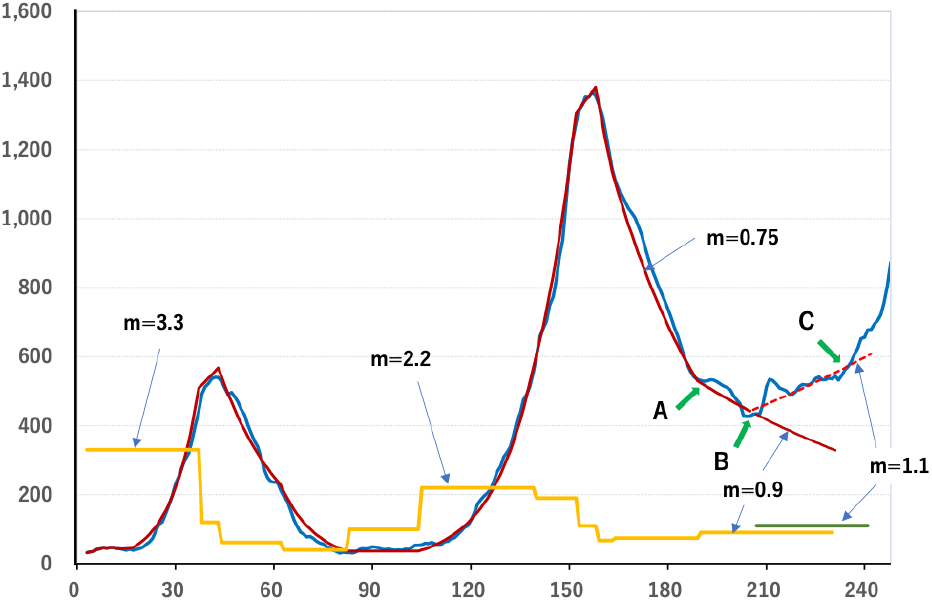
The method of determining time-interval reproduction number *m*(*t*) from the observed infection curve is shown here using the Japanese data of the first and second waves starting at 2020/3/1, as an example [2]. The *x*-axis shows the days after the start and the *y*-axis is the number of infected persons per day. Blue curve is daily infection data averaged by seven days, red curve is calculated best fit curve with *m* = 0.9 within the time interval between A and B, that deviates, however, from the observation beyond B. Then we readjust *m* to be *m* = 1.1 so as to best fit the observed data as shown by the dashed curve. Thus obtained series of reproduction numbers *m*(*t*) are shown by the yellow step line.

Thus, for each time interval, a constant reproduction number is obtained. This number *m* reflects infectivity in a static environment around the virus such as nation’s lifestyle habit, but also influenced by the governmental regulation or behavior change of the public etc. This is different from effective reproduction number in SIR model [4, 5], where *m* is modified by a factor *B*(*t*)*/A* and basically represents only a logarithmic slope of the infection curve.

The startup phase of the first wave where people are not prepared against infection, virus infectivity is strongest, and is defined as basic reproduction number *m*_0_. Unless the virus is replaced by the variant, intrinsic nature is maintained during its multiple waves, and infectivity represented by *m*_0_ appear if virus environment goes back to the stage before the corona.

## 3. Application of the RN-EXCEL model

### 3.1. Infectivity variation and COVID-19 development

By the method developed in II.C above, we can compare the infectivity of each wave by the *m* value. For instance, Fig. 4a shows the daily infected persons in the case of Japan from the 1st wave until the 8th wave of omicron (2020/3/1 until 2022/12/28). Typical reproduction numbers in buildup phases are also shown by the numbers beside the arrows. The same data is shown in logarithmic scale in y-axis in Fig. 4b, with a detailed time-interval reproduction number *m* in y2 axis (right hand side) in linear scale.

**Figure 4:**
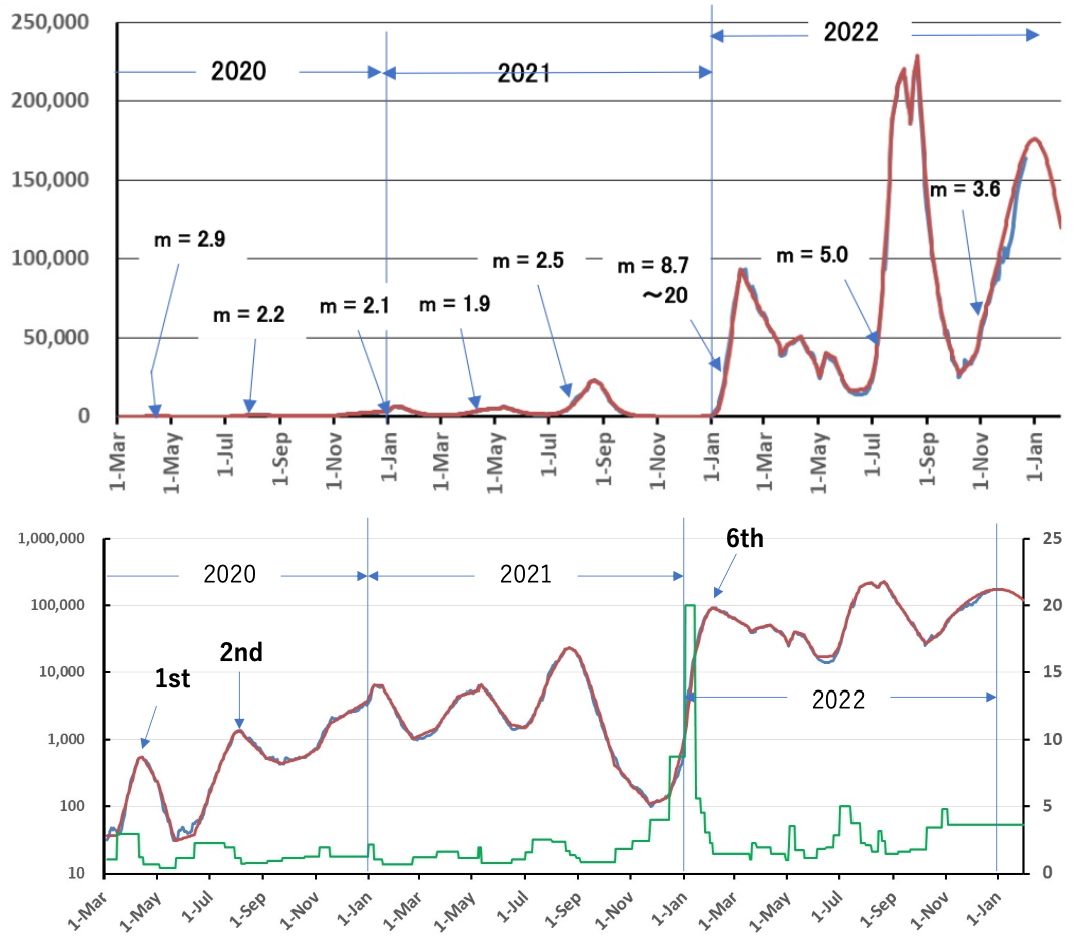
a) COVID-19 waves have been analyzed with time-interval reproduction number for the Japanese case. The y-axis shows daily infected persons in linear scale. The red curve is the calculated best fit results, which agree with the observation shown by blue curve. b) Logarithmic expression of the daily infected persons in left y1-axis, and the corresponding time-interval reproduction number in right y2-axis. The green curve denotes the time-interval reproduction number *m*.

In Fig. 4a and b, we observe that relatively high infectivity for the first and second waves, in spite of their low wave peaks, and that high values of *m* by the omicron variant, in particular, high *m* value of 8.7 (for a time-interval of 2021/12/16 *−*2022/1/2) reaching up to 20 (for a time-interval of 2022/1/3 *−* 2022/1/14) at the buildup phase of the omicron sixth wave. These extremely high values, when averaged over two time-intervals, gives an average value of *m*=12.2.

The other observation is that at earlier phase of infection like the first and second waves, *m* at the peak is *∼*1.0 from Eq. (3) because *m*_*eff*_ = 1.0 and *B*(*t*) *∼ A*, but two years later, at the sixth wave, the *m* value at the peak is *m ∼* 1.4. This is due to the shielding effect by the immune holders, as mentioned earlier.

In general, the time-interval reproduction number *m* changes subject to governmental regulations or to the self-protective behavior change of the people. We find in most cases of repetitive waves, the reproduction number *m* at the buildup phase becomes somewhat smaller for the 2nd and 3rd wave, compared with the 1st wave, if the virus variant is the same. Even if no governmental regulation is applied for the 2nd or 3rd wave, people are prepared to protect themselves against infection more cautiously than the first attack. These phenomena are often seen in many countries and provinces.

We now have a complete set of observed data base, but we emphasize that occasional antibody tests are necessary to know the time dependence of *α*. In conjunction with asymptomatic infected persons with vaccination, attention must be payed in the model that vaccination to these people means an unavoidable waste of injection because they already have immunity. To make a predictive calculation, *m* value need to be known. Three approaches are practically valuable:

1. When nothing is changed in the environment of the virus, *m* stays at a same value. This is the case if governments do not change regulation, and citizens are expected to spend without behavior change. Then, the same *m* value can be extended to the future.
2. After several experiences of the infection waves, we come to know typical *m* values corresponding to each social environment. Choose appropriate *m* value to make predictive calculation under the similar environment foreseen.
3. When the government deregulates, and the public behavior change is expected to return to the one before the corona, then infection will follow a virus infectivity characterized by the basic reproduction number *m*_0_. The end of infections can be obtained only when no increase of infections is predicted with *m*_0_. This is very important to distinguish between a temporal equilibrium and the end of infection.

In the following, we want to show some actual cases from Japan, UK. and USA., as examples.

#### Japan

Fig. 5 shows the case of Japan 3rd wave build up. The observed *m* number from the bottom of the infection curve, increased gradually from *m* = 1.1, 1.25, and 1.9 up to the 250th day counted from the 1st day (2020/3/1). The last fitting time-interval is indicated by red allow (*x* =251-257) with *m* value of 1.9.

**Figure 5:**
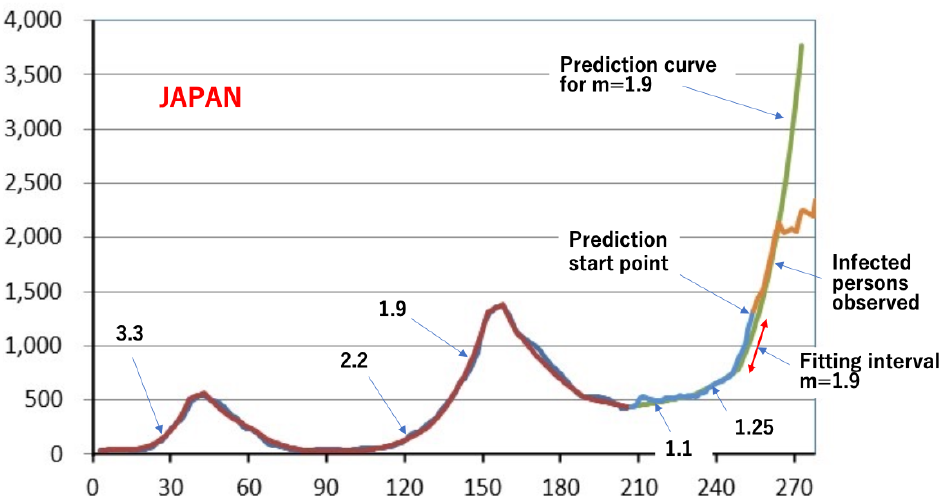
Predicted Japanese 3rd wave buildup phase. The x-axis is the days from the beginning of COVID-19 (2020/3/1), and y-axis shows daily infected persons.

Based on this observed build up phase (blue color), a curve was extended for *m* =1.9 (yellow-green curve) for prediction. Beyond the point shown by “prediction start point”, an orange-colored curve indicates actual observed data after the start of prediction. We see observed infected number of persons exactly follow this curve, but deviation started about 10 days after the start of prediction. The Japanese government regulated people travels by an alert in the 1st wave, but for the buildup of the 3rd wave, no such actions were taken except for repeatedly asking people to avoid close contacts. Therefore, the deviation from the prediction curve is due to self-defense behavior change of people knowing sharp increase of infection, especially at a time daily infection rate exceeding the peak of the former wave.

#### United Kingdom

In UK, a similar trend was observed as shown in Fig. 6. Fitting zone was 2022/8/25*−* 9/22 with *m*=2.37 and prediction started hereafter. The red curve is the calculated result and the green curve is observation. Just from the start of prediction, the UK government started regulation, and strengthen regulation about three weeks later (10/14 incl. local lockdown). Soon after it, the observed data (blue line) started to deviate from the *m*=2.37 prediction curve (red curve). Since infection still stayed at a high level exceeding 20,000 persons per day, the UK government started a national lockdown regulation (11/5*∼*) [6]. Then the infection started to decay at about 2 weeks later.

**Figure 6:**
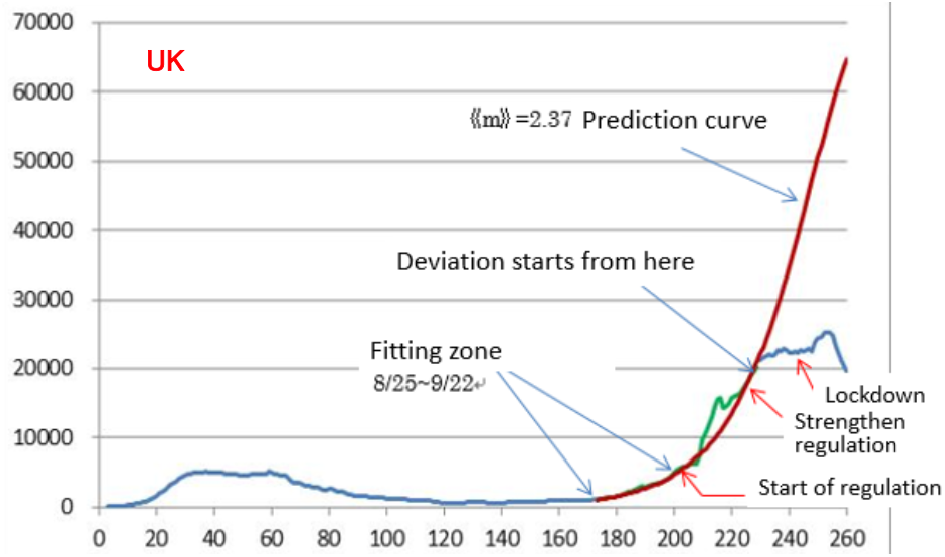
In case of United Kingdom, *m*=2.37 is derived from the data between 2022/8/25 *−* 9/22 shown by the fitting zone and extended for calculation. The observed data are shown by green curve and blue curve. The dates for the governmental regulation were: Start of soft regulation (9/22), Strengthen regulation but by the local government level (10/14), and UK 2nd national lockdown (11/5*−*12/2).

The infection curve in UK reflects the relation between public behavior change and governmental regulation. If no action took place, the infection would have followed *m*=2.37 curve.

#### USA

In USA, Fig. 7 shows the infection curve with a typical *m* value. Compared with other waves, it is sharply peaked at the first attack of omicron wave. At its buildup phase, the infectivity is *m*=6.5, higher than delta variant. Before arriving at the results shown in Fig. 7a, we encountered a difficulty that prediction curve cannot follow observed infection data even though we assumed very high values of *m*=16 *∼* 35 at the end of the 2nd omicron wave as shown in Fig. 7b. It means a huge lack of non-immune holders, susceptible for infection, suggesting more basic problems exist. Since time decay of mutation by the vaccination has already been taken account [7, 8, 9] the residual possibility is reinfection [11, 12].

**Figure 7:**
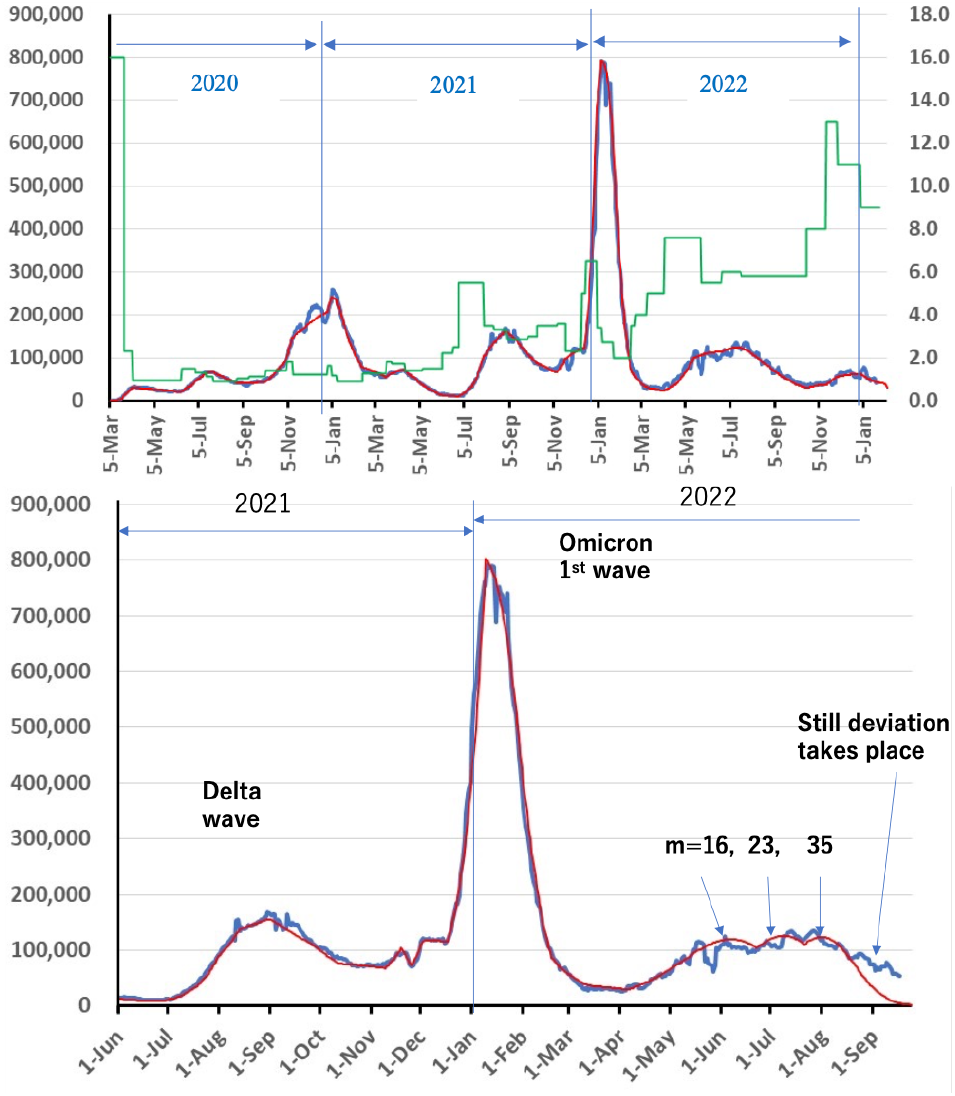
a) Daily infection curve for the Covid-19 virus of the USA in the time period of 2020/3/5 *−* 2023/1/23. The typical time interval reproduction numbers are also shown in green curve with the scale in the y2-axis. b) Deviation of the calculated curve from the observation is significant. To solve this difficulty, assumption of time decay of infection immunities has been incorporated in the calculation. This change significantly relaxed the unreasonably high reproduction number in the 2nd omicron wave as shown in Fig. 7a.

In UK HSA report, direct reinfection data from a cohort study on NHS healthcare workers provides a clear sign of occurrence of reinfection [13], but we want to know for prediction purpose, a degree of conversion from the infected immune holders to non-immune holders in the background of reinfection. Nature News [14] suggests such a possibility by the decay of infectious mutation in a time scale of a year in contrast to the decay of vaccinated immunity in several months. Although the analysis of data is from pre-Omicron variant, if immunity by pre-Omicron infection will also fade away for Omicron, this mechanism would produce a lot of non-immune holders and supply infectious persons.

To check such a possibility, we use a pinpoint data that about 20% of cumulative infectious immunity holders convert to non-immunity holders at one year after former infection. It is particularly important for USA and UK or many other European countries, where a large scale infection took place toward the end of 2020. Since that time almost two years has passed which is enough for infected immunity to fade down to a level less than a half. The result of calculation taking immunity decay is shown in Fig. 7a. In this figure, the infectivity of the second peak is higher than the first peak, but this is explained by emerging lineage EB.5 at this timing. The height of the 1st and the 2nd wave of omicrons is a big difference, but such a trend took place in approaching a herd immunity threshold, since non-immunity rate *B*(*t*)*/A* changes significantly after a large 1st omicron infection wave.

While most of the difficulty are resolved, there still exist one difficulty at the end of Fig. 7a, the third wave of omicron. A large number *m*=13 is still necessary to follow the observed continuing data. This suggests whether emerging omicron lineage such as XBB.1.5 has stronger infectivity [16] or more conversion took place from infectious immunity to non-immunity holders, but this is postponed to a future work. More detailed predictive calculation should also include total time dependent immunity decay curve.

In USA, without taking the background immunity conversion for reinfection into account, calculation did not provide sufficient infectious persons to follow observed data, and hence it is essential to incorporate immunity decay of infection.

### 3.2. Prediction toward the end of infection

As seen in Eq. (4), the next day infection is determined by the effective reproduction number that is modified from the reproduction number by a ratio of non-immune holders to the total population *B*(*t*)*/A*. Let us call this reduction, a “herd immunity effect” where the important driver of this effect is *R*(*t*)*/A*(= (1 *− B*(*t*)*/A*)), an immune holders’ ratio. Through this relation, for a virus of the same infectivity, the spread speed becomes milder with a progress of infection owing to an increase of *R*(*t*)*/A*. Therefore, for predictive calculation, *R*(*t*)*/A* is a key variant that should be monitored continuously. Many countries in US, Europe, India, South Africa have been analyzed by the RN-EXCEL model, and most of these countries are now close to the herd immunity threshold.

Here, we pick up Japanese case since it has produced small number of infected persons, and most of immunity holders are from vaccination. Therefore, a ratio of immune holders in the population is much smaller than those of other countries. Fig. 8 shows observed infected number of persons and the calculated fitting curve up to 2022/11/14 and nearby prediction for the 8th wave in Japan. At the beginning of the 8th wave, the time-interval reproduction number was *m* =4.8, but soon after, the buildup speed was relaxed to *m* = 3.6 for a time-interval of 2022/11/4 *−* 11/14 shown in the Fig. 8. Beyond 2022/11/15 is the prediction calculation assuming the same reproduction number. If this assumption holds, the infection will soon reach a peak and start waning. However, note that the 8th peak does not mean herd immunity threshold for omicron. This peak is a herd immunity threshold for *m* = 3.6, and not for the basic reproduction number *m* = 12.2. For instance, at the tail of the 8th wave when the infection is comparable to the bottom after the 7th wave, about 22.6million (about 18% of Japanese population) of non-immunity holders still exists.

**Figure 8:**
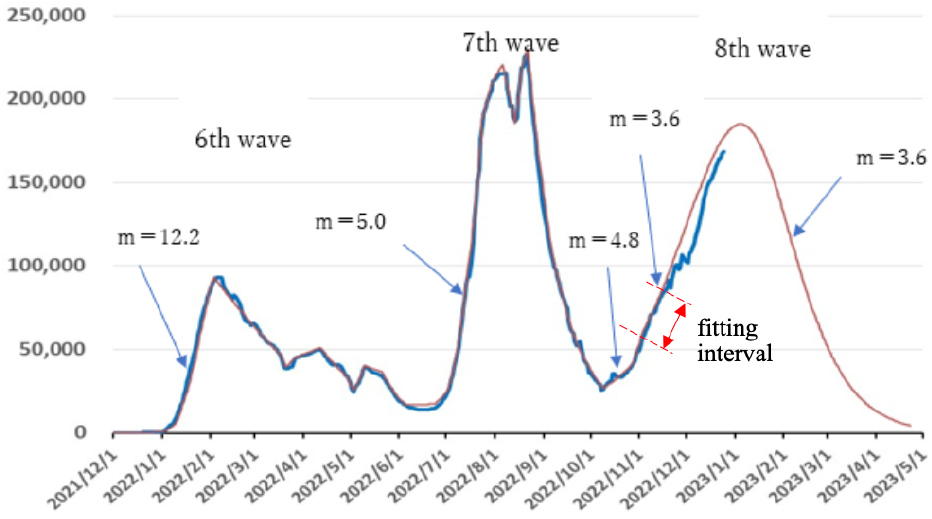
Observed daily infected persons in Japan, and predicted infection curve for the 8th wave.

Therefore, if the virus environment is changed just as before the COVID-19 (before 2020/3/1) world, the infection spreads again as in Fig. 9 with a reproduction number *m*=12.2. Here, we use the 71% of effectiveness of omicron specific vaccination reported by the Niid on 2022/12/13 [17] and assumed no time dependent decay of effectiveness in contrast to the former vaccinations.

**Figure 9:**
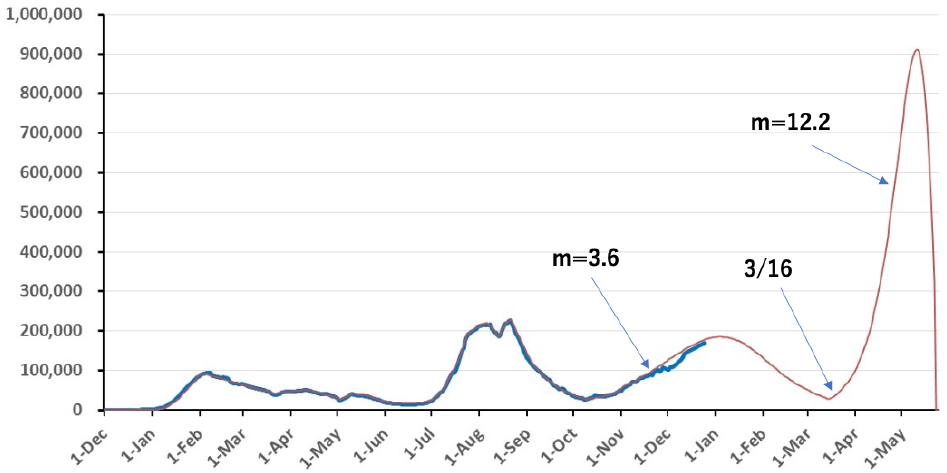
On 2023/3/16, the infected person is below the bottom after the 7th wave. If any governmental regulations or human behavior change are dissolved just as before the corona world, then the omicron virus starts to build-up at the reproduction number *m*=12.2.

In Fig. 9, the peak reaches to 900 thousand persons per day which is a real herd immunity threshold for *m*=12.2 omicron virus. This may not take place, since (if infection spreads too fast), people try to change their behavior that plays to control infection spreads even without governmental regulations. Therefore, a peak will be suppressed like ones of 6th and 7th wave (where no governmental regulation had been set). However, it simply means that infection deferred. Important is a rate of the residual non-immune holders, that indicates a potential of the forthcoming waves. To finalize omicron infection, while the peak had to be suppressed below some level, for instance, at a level comparable to the 8th wave, one way to control is to increase *m* value from the present one to the final herd immunity reproduction number continuously. This approach means the regulation including behavior change gradually return to the state of before the corona world, namely, before 2020/3 in case of Japan. Shown in Fig. 10, *m* is changed from 3.6 to 12.2 continuously.

**Figure 10:**
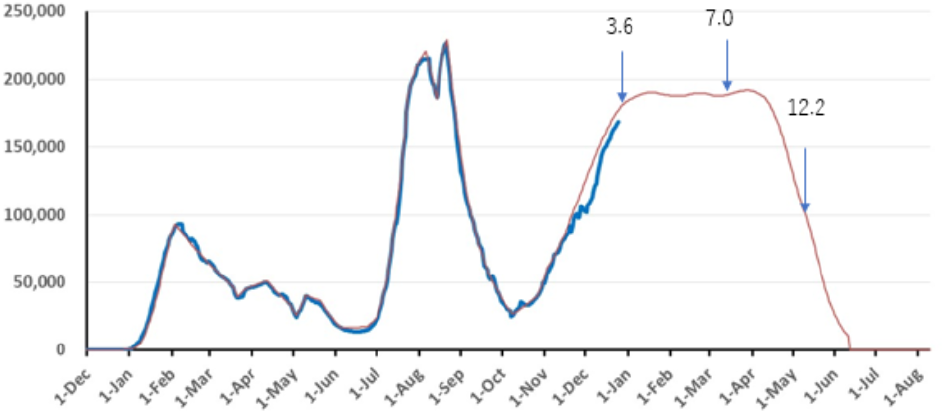
If *m* is changed from 3.6 to 12.2 continuously, the prediction curve can be formed like a plateau shape. The herd immunity threshold can be met during a decay slope and the final end of the infection is achieved about a half year later.

Time changing balance of the immune holders distribution corresponding to Fig. 10 can be seen in Fig. 11. Since time decay of vaccine effectiveness is significant, recovery by the booster or new omicron vaccine is modest. The difference between blue and green curves is the contribution by the infectious immunity. Therefore, for the total effective immune holders to reach a level compatible to the herd immunity threshold population rate, the immunity by infection must be increased significantly. During the plateau period in Fig. 10, this could be met. It is also shown that almost no or very few non-immune holders are left at the end of infection due to high infectivity of omicron variant.

**Figure 11:**
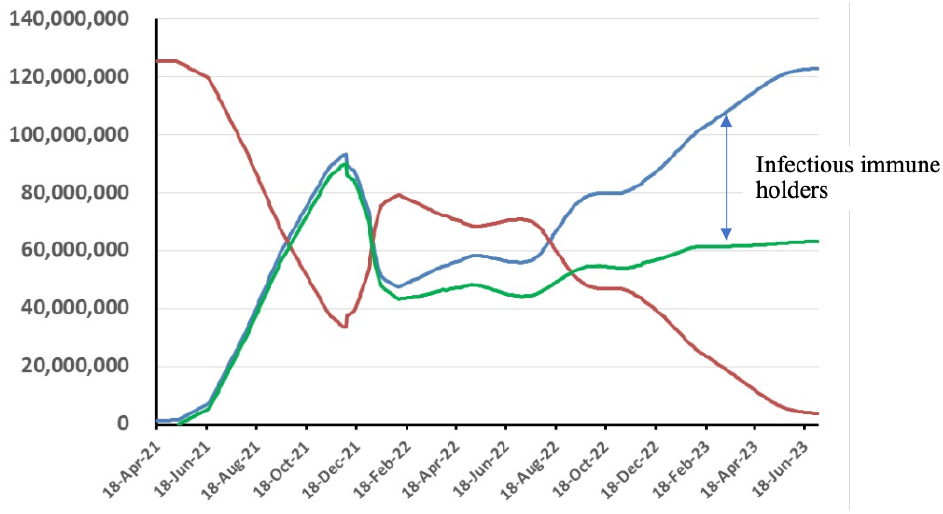
Time variation of the immunity sharing of the Japanese population in the case of Fig. 10. Red: Non-immunity holder, Green: effective vaccine immune holder, Blue: total immune holder. The difference between blue and green indicates infected immunity holders.

Thus, on the basis of reasonably assumed effective vaccination speed and *m* numbers, we can make predictive calculations using RN-EXCEL model. We have done a lot of analysis using the RN-EXCEL model applying world-wide, to various countries such as Japan, UK, USA, India, South Africa, and many other European countries. Particularly, owing to available continuous informations, we have followed the cases in Japan, UK, and USA. Comparing these countries, the status of Japan is quite different in a state of total immunity ratio to the population. This is due to a low generation of the infected persons in the first half of COVID-19 infection. Since vaccine immunity will not be easily increased due to a time decay of effectiveness, the total immunity percentage is modest as seen in Fig. 11, and the distance to the herd immunity threshold is quite far. The required immunity level for omicron variant to achieve herd immunity threshold is close to 90%, or more depending upon the country. The UK and USA already seems to have achieved immunity threshold, India and South Africa too owing to a high asymptomatic infectious percentage, but Japan is still in the process of increasing infectious immunity holders.

## 4. Summary

The RN-EXCEL Model has been proposed for practical use of infection such as COVID-19 pandemic. Features of the Reproduction Number Excel Model are:

1. The model is structured to base only on observed data and herd immunity concept.
2. Physical picture is clear by introducing a concept of time-interval reproduction number for the base of index function. This makes possible to analyze breaking factor such as governmental regulation, public behavior change, and to incorporate time-dependent vaccination and its efficacy, reduction of vaccine effectiveness etc. for which traditional SIR model is difficult to handle.
3. Asymptomatic and mild infected persons that are not counted by the PCR tests have been estimated statistically by the aid of antibody tests. By this all the data are based on the observation.
4. Total number of immune holders is always monitored to know a distance from the present situation to the herd immunity threshold and to the end of infection.

These features are combined in the RN-EXCEL model, and after short learning period, it can provide prompt practical predictions required for political judgements.

The authors wish this simple and practical RN-EXCEL model be used anywhere in the world for infectious disease that may appear in future.

## Data Availability

All the data in this article are publically available.

## Appendix: SIR model vs. RN-EXCEL model

The famous infection model is the SIR model. The SIR model is based on a coupled differential equation for numbers of susceptive persons *S*, infectious persons *I* and removed persons *R*. These numbers are related each other as

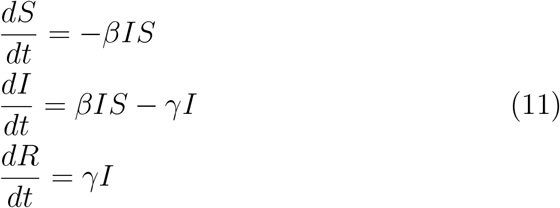

The total number *A* is the sum of these three states *A* = *S* + *I* + *R*. The infections persons are created when infectious persons (*I*) meat with susceptive persons (*S*) with the strength rate of *β*. The infectious persons then are recovered or died by the disease and eventually removed (*R*) from either *I* or *S* groups with the rate *γ*. The difficulty of the SIR model to apply to the actual case as COVID-19 is the infection strength *β* changes frequently with time due to the governmental policy as lock-down and/or mental pressure of individuals. In addition, we have to include vaccination to change the status of *S* to *R*. Additionally the immune stage *R* is not really to be removed, because the immune effect is weakened as time elapses. All these effects make the actual SIR model highly complicated.

In the time before the vaccination was introduced, the original SIR model could be used as given in Eq. (11). In the SIR model, we ought to change *β* frequently so as to reproduce the daily incidence of infectious persons 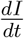, which were announced daily. This is possible, but quite cumbersome. After the vaccination was introduced, there should be a term to remove *S* and increase *R* with a rate of vaccination, which was again highly time dependent. In addition, the effect of the vaccination decreases with time, and there should be a term to reduce *R* and increase *S*. Hence, after the vaccination was introduced, we had to change the rate of vaccination announced frequently.

In the RN-EXCEL model, all the essence of the SIR model was taken into account with the concept that the daily announcement of the incidence of the infectious persons 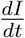 was used as input so as to follow the development of COVID-19 at the daily basis. Hence, we had to take the time step as 1 day and write the SIR equation in the difference equation form. Furthermore for the daily change of the incidence, the important quantity to be fixed daily was the multiplication factor *m* using Eq. (2). There are several unique features in the RN-EXCEL model. The effect of *γ* is introduced as the duration *τ* in the RN-EXCEL model so as to take into account the cure/death effect at the daily basis. Since the essence of the infectious disease is the herd immune effect, the RN-EXCEL model divide the entire population into two groups (*B*: non-immune group, *R*: immune group) and the introduction of the vaccination and its prevention-deterioration rate in a simple manner. All these modeling ideas made the infection development easy to handle using the EXCEL sheet, and made possible the analysis of complicated changes of various effects from the beginning until its end in various countries.

## Acknowledgement

The authors wish to express their sincere appreciation to Prof. Shigetaka Katow, eminent infectious-disease researcher, for his understanding of the model, valuable comments, and continued supports.

## Notes

### Competing Interest Statement

The authors have declared no competing interest.

### Funding Statement

This study did not receive any funding.

### Author Declarations

Infection data used in this article are mostly from Nikkei world map for corona infection which are collected by Johns Hopkins University in the U.S., and the data of infection in Japan are from NHK corona site collected by Ministry of Health, Labor Standards.

